# Predicting Antiseizure Medication Outcomes in Early Diagnosed Epilepsy: A Multimodal Framework Using EEG, MRI, and Clinical Data

**DOI:** 10.1101/2025.03.12.25323644

**Authors:** Duong Nhu, Jiahe Liu, Richard Shek-kwan Chang, Daniel Thom, Zhibin Chen, Mohamad Nazem-Zadeh, Alison Anderson, Sarah Barnard, Jacqueline French, Patrick Kwan, Zongyuan Ge

## Abstract

Accurate prediction of antiseizure medication (ASM) outcomes is crucial for optimising epilepsy treatment. We propose a multi-modal deep learning framework that integrates electroencephalography (EEG), magnetic resonance imaging (MRI), clinical factors, and molecular drug features to enhance ASM outcome prediction. Our approach includes EEG Q-Net, a pre-trained quantisation model capturing finegrained temporal EEG patterns, and MRI embedding from BiomedCLIP. To capture chemical similarities and interactions for the ASMs, we employed a pre-trained model trained on thousands of medications to generate embedding for SMILES (Simplified Molecular Input Line Entry System). Our findings underscore the benefits of multimodal integration for personalised epilepsy management, as our fusion model achieved an AUC of 0.75, an improvement over the best unimodal model (0.71).

## 1 Introduction

Epilepsy is a complex spectrum of disorders that affects around 50 million people worldwide [23]. It is characterised by recurrent unprovoked seizures that differ in type, cause (including brain structural, genetic, metabolic or autoimmune), and severity. Uncontrolled seizures can cause substantial health, psychosocial, and economic impacts to patients, including increased hospitalisation [8], comorbidities, premature mortality, and decreased quality of life [3]. The mainstay of treatment for epilepsy is anti-seizure medications (ASMs) which suppress seizure occurrence without modifying the underlying disease process. Current ASMs fail to control seizures in one-third of patients [2], largely due to our limited understanding of the complex neurological mechanisms underlying epilepsy and individual responses to treatment [22]. Finding the right ASM(s) for an individual has remained a largely trial-and-error process of selecting from more than 25 different drugs with critical time lost in trying ineffective ones until the ‘right’ one is found. Patients have variable responses to ASMs, including serious adverse effects such as hypersensitivities, and there are major barriers in diagnostic methods to improve ASM safety (e.g. expensive lab-based testing for genetic markers). There is also a lack of non-invasive methods to accurately monitor seizures in the community. The consequences of these unmet needs are years of reduced quality of life, lost productivity, and increased mortality.

With the success of data-driven advancement in many fields in recent years, there have been efforts to develop AI-based approaches for predicting ASM outcomes and personalised treatment [10,5]. These studies only used clinical factors that did not cover the whole epilepsy spectrum, resulting in low performance. de Jong [5] extended these studies by including genomic information, achieving reasonable results. However, whole genome sequencing is expensive and adds additional cost to patient care [6], and is not always covered by national health insurance. In comparison, as a standard protocol, most patients with epilepsy undergo either or both electroencephalogram (EEG) recordings and magnetic resonance imaging (MRI) scans for diagnosis [11]. Therefore, predicting ASM outcomes with existing EEG and MRI may be more cost-effective.

Although raw EEGs and MRIs carry drug-resistant epilepsy (DRE) characteristics [26,20], they remain underutilised in AI models. EEG microstates [26] and functional connectivity [21] demonstrate differences between drug-responsive and DRE patients. MRI-detected lesions [20] are linked to DRE; however, they are not always shown in standard MRIs [4]. Recent deep learning models using raw MRIs have shown promise in predicting neurological outcomes [19], suggesting the potential for ASM response prediction through automatic feature extraction.

Moreover, existing works in ASM outcome prediction only used one-hot encoding for available drugs [10]. This limited the performance and the generalisability of the model to the included ASMs. There have been works [14,15] developing deep learning with large corpora of medications to learn the embedding for SMILES (Simplified Molecular Input Line Entry System). SMILES encodes drug structures as character sequences (e.g. ‘CC(=O)N1CCCC1C2CCOC2’ for levetiracetam, Figure 1(a)). SMILES embeddings capture chemical relationships between drugs, allowing testing on any ASM, including unseen ones, and enhancing performance for rare ASMs.

**Fig. 1.**
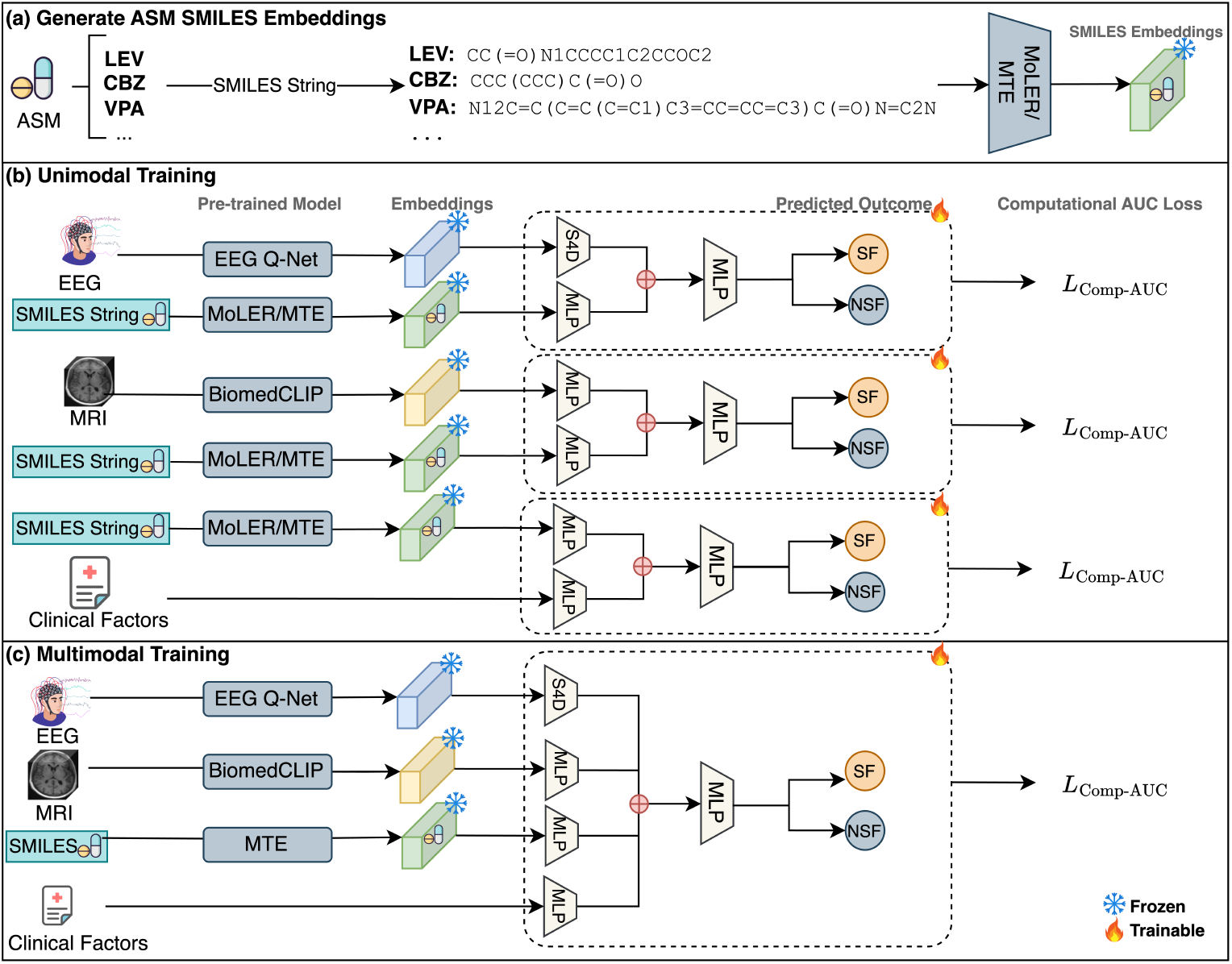
The pipeline of our proposed framework. **(a) SMILES Embeddings Generation:** SMILES strings for ASMs were converted into embeddings using two pretrained models: MoLER [14] and Molecular Transformer Embeddings (MTE) [15]. **(b) Unimodal Training:** Each modality (EEG, MRI, and clinical factors) was trained separately with SMILES embeddings. EEG and MRI embeddings were extracted using EEG Q-Net and BiomedCLIP [25], respectively. **(c) Multimodal Training:** The best-performing embeddings were concatenated and passed to an MLP fusion layer.

This project aims to develop a robust system to predict first ASM monotherapy outcomes using a multimodal deep learning approach that integrates clinical data, EEG, MRI, and SMILES embeddings. These routinely collected data ensure the model is easily translatable to clinical practice for personalised care.

## 2 Methods

The objective is to predict the chance of seizure freedom when a patient is administered an ASM. We considered this a binary classification task by encoding the outcomes as 1 (seizure-free) and 0 (not seizure-free). The inputs to the model will always contain SMILES embedding, with additional modalities depending on the specific setup. The output of the model will be the probability of seizure freedom if the patient takes a particular ASM as demonstrated. Our framework is demonstrated in Fig. 1.

### 2.1 EEG Pre-training Framework

Modelling continuous signals like EEG is challenging due to infinite variability across patients and devices, limiting generalisability [16,17]. Recent deep learning methods tackle this by discretising signals with product quantisation that maps them to discrete codebooks [1,12]. Structured State Space models (S4) also show promise by quantising signals through bilinear transformation [9]. To address this, we developed EEG Q-Net, a pre-trained quantisation deep learning architecture for EEG signals (Fig. 2). Our architecture shares similarities with the wav2vec 2.0 architecture [1] but with a few modifications.

**Fig. 2.**
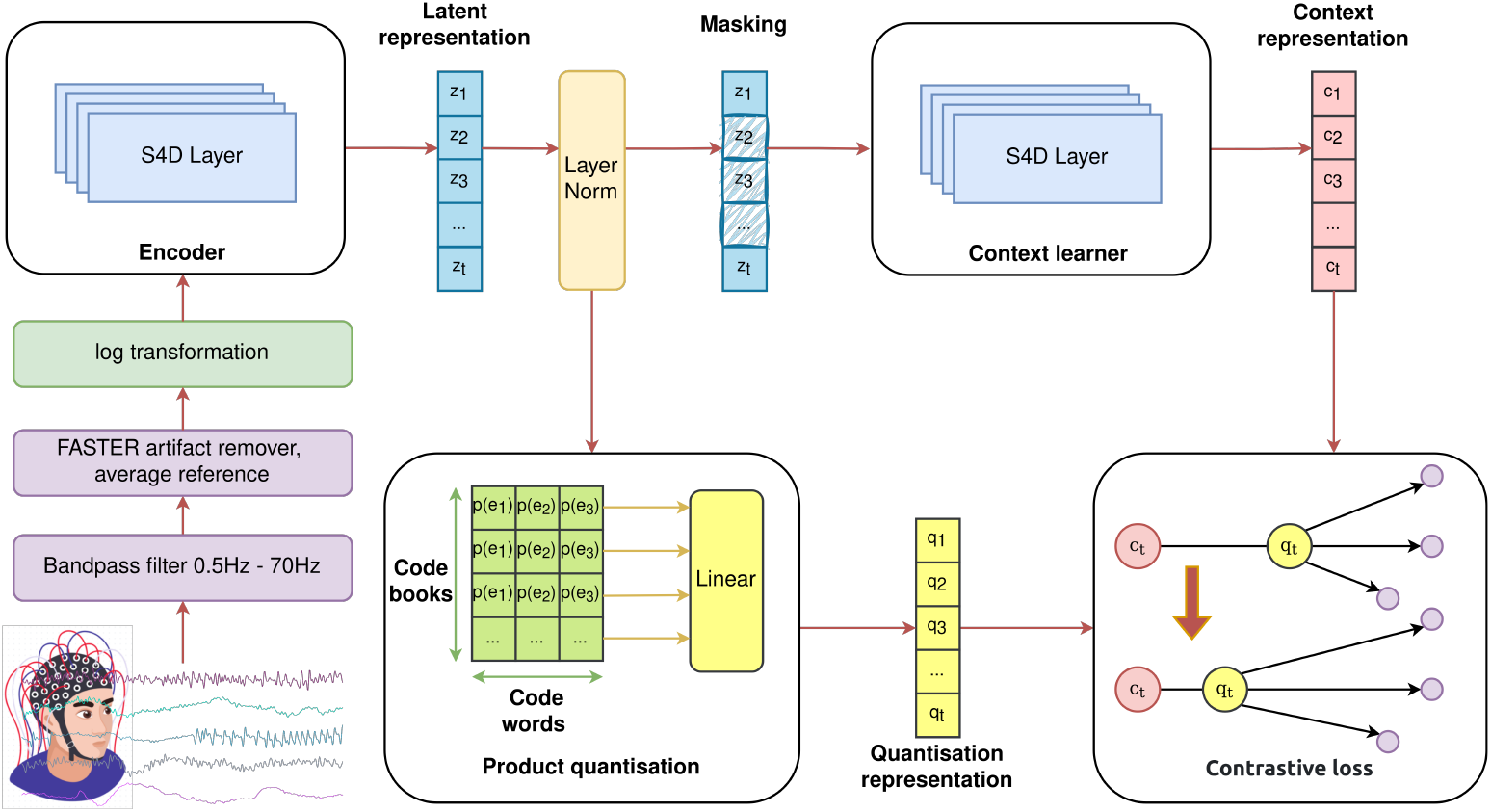
The architecture of our proposed EEG Q-Net.

#### Quantisation Representation

We used product quantisation to map each *z*_*t*_ to a codeword *e*_*t*_ from a codebook *E*^*d*^, where the codebook size d is tunable. The probability *p*(*e*_*t*_) for choosing a codeword *e*_*t*_ was calculated using Gumbel softmax over the whole codebook, with the same decay strategy as in wav2vec 2.0 [1]. The minimum and maximum temperatures were 0.5 and 2, respectively. A linear operation was applied to obtain the quantisation representation *q*_*t*_.

#### Masking

We randomly sampled N timesteps of the latent representation and masked M consecutive steps. We zeroed out the presentation of each masked timestep. In our experiment, we set N=5 and M=10.

#### Context Learner

We used an S4D model to fill the masked latent representation based on the surrounding context, producing context presentation *c*_*t*_ for each timestep.

#### Loss Function

For masked timestep *t*, the model needs to learn *c*_*t*_ such that *c*_*t*_ is as close to *c*_*t*_ as possible. We define our contrastive loss function *L*_*m*_ as a normalised temperature-scaled cross-entropy loss, where the temperature was 0.1, and the similarity metric was cosine similarity. To avoid the pitfall where the model only chooses a small set of codewords, we added a diverse regularisation *L*_*d*_ that is the softmax distribution of the codeword 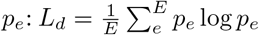. The final loss function is: *L* = *L*_*m*_ + *αL*_*d*_, where *α* was set to 0.2 in our experiments.

#### EEG Embedding

We compared two EEG embeddings: (1) normalised latent representation and (2) context embedding. Each timestep has an embedding, averaged over a window, which was then concatenated to form the full EEG embedding. Shorter EEGs (<20 min) are padded with the last window.

We used 10 codebooks with 18 codewords each. The encoder and context learner each had 16 S4D blocks (state size 512, dimension 128), with the encoder downsampling signals every two blocks. For negative samples, 200 timesteps were randomly selected from the minibatch. We used the AdamW optimiser with a learning rate of 1e-4, weight decay of 0.1, and cosine annealing with warm restarts every 20 epochs.

### 2.2 ASM - SMILES Embedding

We extracted the SMILES notation for each ASM from Drugbank, https://go.drugbank.com/. Two embeddings were tested: MoLER [14], a graph-based model accounting for scaffolds and bonds, and Molecular Transformer Embeddings (MTE) [15], an end-to-end transformer encoder. For MTE, the final embedding was obtained by averaging the character embeddings.

### 2.3 MRI Feature Extraction

Foundation models have been shown to be powerful pre-trained models that demonstrate effective transfer learning, particularly for tasks with limited data [25]. In this study, we employed BiomedCLIP [25], a vision-language foundation model trained on a large corpus of MRIs, to extract features (512-dimensional) from 2D MRI scans. Recognising that axial views alone are insufficient for epilepsy diagnosis, especially refractory epilepsy [13], we compared two strategies: (1) only axial slices and (2) multi-view stacking all three views, axial, coronal, and sagittal.

### 2.4 Multimodality Fusion Architecture

As shown in Fig. 1, we compared unimodal and multimodal models to evaluate the impact of integrating multiple data modalities on ASM outcome prediction. SMILES embeddings (512-dimensional vectors from MoLER or MTE) were combined with EEG, MRI, and clinical factors in separate unimodal baselines. A multilayer perceptron (MLP) was used to fuse concatenated outputs from domain-specific MLPs. Each MLP consists of three blocks, comprising a linear layer, ReLU activation, dropout, and layer normalisation. EEG features were extracted using an S4D network (8 layers). The best-performing embeddings were then integrated into the final multimodal model.

We directly optimised the AUC by using the compositional AUC loss with Primal-Dual Stochastic Compositional Adaptive (PDSCA) optimiser [24]. We applied a cosine annealing schedule to both the outer and inner learning rates. The number of inner steps was set to 2, with both learning rates of 0.05 and a weight decay of 0.1. To mitigate class imbalance, we ensured each minibatch (16 samples) contained an equal number of seizure-free and not seizure-free samples

## 3 Experimental Setup

### 3.1 Datasets

We collected EEG, MRI, and clinical data from the Human Epilepsy Project (HEP1) and the Alfred Health Hospital in Australia (Alfred). HEP1, http://www.humanepilepsyproject.org, is an international multicentre observational study between 2012 and 2020 that recruited participants within four months of starting ASM for newly diagnosed focal epilepsy. Approval for HEP1 was obtained centrally from the New York University Institutional Review Board and locally from the Institutional Research Ethics Boards of each participating site. The Alfred dataset included patients diagnosed between 2018 and 2022 and approved by the Alfred Health Ethics Committee. These patients were administered first-line ASMs, such as levetiracetam, carbamazepine, and valproate. There were seven drugs in our dataset. Seizure freedom was defined as no seizures for 12 months on initial ASM monotherapy, while treatment failure involved therapy changes within this period. Overall, we obtained 85 seizure-free and 272 not-seizure-free patients, including generalised and focal epilepsy. EEG recordings and MRI scans were routinely collected before medication administration.

### 3.2 Data Pre-processing

#### EEG

We extracted the first 20 minutes of recordings, excluding the initial five to remove calibration artifacts, hypothesising that this duration captures stable functional connectivity sufficient for treatment outcome prediction. Recordings shorter than 20 minutes were discarded. The signals were segmented into 10-second windows, downsampled to 256 Hz, and filtered (0.5–70 Hz) before undergoing artifact removal via FASTER [18], average referencing, and then log-normalised. All EEGs used the 10-20 system. We only kept 19 electrodes, excluding the ear channels as many EEGs did not record these.

#### Clinical Factors

We collected 16 key variables linked to ASM outcomes [10], including sex, age (categorised as <18, 18–29, 29–46, >46), alcohol abuse, and psychological disorders. We categorised these values as integers, starting from 1, with missing data as 0.

#### MRI

We focused on T1-weighted scans, standardising them to RAS+ orientation and extracting middle slices. Each slice underwent min-max normalisation (0–255 intensity range) and rotation for consistency, followed by brightness, contrast, and sharpness enhancements to improve structural visibility. The preprocessed images were then fed into a pre-trained MRI deep learning model for feature extraction.

### 3.3 Evaluation Metrics

We used 5-fold cross-validation (CV) to assess the generalisability. We primarily optimised the area under the receiver operating characteristic curve (AUC) and reported the average results. Additionally, we measured sensitivity (Sens), specificity (Spec), precision (Prec), and the area under the precision-recall curve (AUPRC), with thresholds set to maximise balanced accuracy on the train set.

### 3.4 Implementation Details

For the EEG Q-Net, the training ran for at least 100 epochs, stopping if the loss did not improve for 10 epochs. For the unimodal and multimodal models, we trained the model for at least 50 epochs and stopped when the validation AUC did not improve for 10 epochs. We implemented our models with the PyTorch Lightning framework [7] on an NVIDIA A100-40GB GPU.

## 4 Results

### 4.1 Unimodal Results

Table 1 summarises the unimodal results. The base models using clinical factors with SMILES embeddings achieved similar average 5-fold CV AUCs for both MoLER and MTE (0.71, standard deviation-SD: 0.06 and 0.05, respectively), outperforming the ASM one-hot encoding (AUC: 0.68, SD: 0.02). For EEG, the normalised latent embedding achieved AUCs of 0.64 (SD: 0.05 and 0.02). In terms of MRI, the BiomedCLIP embeddings of the MRI multi-view had the highest AUC of 0.69 (SD: 0.06). Although EEG and MRI models had lower AUCs than the clinical-factor models, the best EEG models had higher precision.

**Table 1.** 5-fold CV Average Results of Unimodal Models.

| **Clinical Factors + SMILES** |  |  |  |  |  |  |
| --- | --- | --- | --- | --- | --- | --- |
|  | SMILES<br>Embedding | Sens<br>(SD) | Spec<br>(SD) | Prec<br>(SD) | AUC<br>(SD) | AUPRC<br>(SD) |
| Clinical<br>Factors | MoLER | 0.68 (0.13) | 0.69 (0.10) | 0.41 (0.07) | **0.71 (0.06)** | 0.43 (0.09) |
|  | MTE | 0.65 (0.10) | 0.71 (0.07) | 0.41 (0.04) | **0.71 (0.05)** | 0.44 (0.07) |
|  | One-hot | 0.72 (0.08) | 0.55 (0.13) | 0.33 (0.05) | 0.68 (0.02) | 0.42 (0.07) |
| **EEG + SMILES** |  |  |  |  |  |  |
| EEG<br>Embedding | SMILES<br>Embedding | Sens<br>(SD) | Spec<br>(SD) | Prec<br>(SD) | AUC<br>(SD) | AUPRC<br>(SD) |
| Context | MoLER | 0.39 (0.16) | 0.81 (0.02) | 0.46 (0.01) | 0.63 (0.04) | 0.36 (0.04) |
|  | MTE | 0.35 (0.05) | 0.85 (0.01) | 0.46 (0.08) | 0.64 (0.02) | 0.39 (0.04) |
| Latent | MoLER | 0.31 (0.06) | 0.88 (0.01) | 0.47 (0.03) | **0.64 (0.05)** | 0.39 (0.05) |
|  | MTE | 0.39 (0.24) | 0.79 (0.02) | 0.45 (0.01) | **0.64 (0.02)** | 0.39 (0.04) |
| **MRI + SMILES** |  |  |  |  |  |  |
| MRI View | SMILES<br>embedding | Sens<br>(SD) | Spec<br>(SD) | Prec<br>(SD) | AUC<br>(SD) | AUPRC<br>(SD) |
| Axial | MoLER | 0.63 (0.15) | 0.63 (0.16) | 0.35 (0.08) | 0.65 (0.06) | 0.34 (0.05) |
|  | MTE | 0.50 (0.14) | 0.72 (0.05) | 0.34 (0.09) | 0.67 (0.07) | 0.35 (0.08) |
| Multi-View | MoLER | 0.48 (0.16) | 0.70 (0.12) | 0.32 (0.06) | 0.63 (0.07) | 0.36 (0.06) |
|  | MTE | 0.57 (0.10) | 0.71 (0.08) | 0.37 (0.04) | **0.69 (0.06)** | 0.35 (0.04) |

### 4.2 Multimodal Results

We selected the EEG Q-Net latent embedding, MRI multi-view, and SMILES MTE embeddings for further multimodal experiments as they achieved the best AUC and AUPRC scores for their respective unimodal tests. As shown in Table 2, the MRI model yielded an AUC of 0.71 (SD: 0.06), which is similar to the unimodal clinical model. When EEG embeddings were combined with clinical factors, the AUC improved to 0.74 (SD: 0.07). Finally, integrating all modalities produced the best overall performance with an AUC of 0.75 (SD: 0.07), emphasising the benefits of a multimodal approach.

**Table 2.** 5-fold CV Average Results of Multimodal Models.

| **EEG + Clinical Factors + SMILES** |  |  |  |  |  |  |
| --- | --- | --- | --- | --- | --- | --- |
| EEG<br>Embedding | SMILES<br>Embedding | Sens<br>(SD) | Spec<br>(SD) | Prec<br>(SD) | AUC<br>(SD) | AUPRC<br>(SD) |
| Latent | MoLER | 0.71 (0.07) | 0.68 (0.07) | 0.42 (0.09) | 0.74 (0.07) | 0.46 (0.09) |
|  | MTE | 0.74 (0.07) | 0.62 (0.06) | 0.38 (0.06) | 0.74 (0.07) | 0.47 (0.07) |
| **MRI + Clinical factors + SMILES** |  |  |  |  |  |  |
| MRI View | SMILES<br>Embedding | Sens<br>(SD) | Spec<br>(SD) | Prec<br>(SD) | AUC<br>(SD) | AUPRC<br>(SD) |
| Multi-View | MTE | 0.62 (0.12) | 0.72 (0.09) | 0.39 (0.07) | 0.71 (0.06) | 0.41 (0.07) |
| **EEG + MRI + Clinical factors + SMILES (MTE)** |  |  |  |  |  |  |
| EEG<br>Embedding | MRI View | Sens<br>(SD) | Spec<br>(SD) | Prec<br>(SD) | AUC<br>(SD) | AUPRC<br>(SD) |
| Latent | Multi-View | 0.75 (0.14) | 0.67 (0.07) | 0.41 (0.06) | **0.75 (0.07)** | 0.45 (0.07) |

## 5 Discussion

EEG and MRI unimodal models had lower AUC scores than clinical-factor models. The axial MRI view, when used alone, performed worse than the multi-view approach, suggesting that relying on single-plane imaging may not capture the full extent of structural anomalies associated with DRE—a finding consistent with prior research [13]. BiomedCLIP MRI embeddings showed no statistical improvement over clinical models, suggesting 2D MRI slices are insufficient for ASM outcome prediction. However, integrating all modalities improved the overall performance, highlighting their complementary insights. Our model can be adapted for ASM selection by using its probability output to predict seizure freedom and guide effective treatment.

A key advancement in this study was using SMILES embedding, which outperformed one-hot encoding. Unlike one-hot encoding, which only represents categorical data, SMILES embedding captures the structural complexity of drugs in a rich, continuous feature space, enhancing the model’s ability to detect subtle differences in ASM efficacy.

## 6 Conclusion

This study demonstrates that integrating EEG, MRI, clinical factors, and SMILE embeddings enhances ASM treatment outcome prediction in newly diagnosed epilepsy. By leveraging routine multimodal data, our approach improves early treatment decision-making and lays a foundation for personalised patient care. Future work will focus on specialised 3D MRI models, managing missing modalities, and refining co-embedding techniques to optimise data fusion and enhance prediction accuracy and generalisability.

## Data Availability

Data will be made available on request

## 7 Acknowledgement

There was no targeted funding for this study. The creation of HEP was sponsored by the Epilepsy Study Consortium. Funding for HEP was received from industry, philanthropy, and foundations (UCB, Eisai, Pfizer, Lundbeck, Sunovion, the Andrews Foundation, the Vogelstein Foundation, Finding a Cure for Epilepsy and Seizures [FACES], and Friends of Faces).

